# Evaluating the impact on health outcomes of an event that resulted in a delay in contact tracing of COVID-19 cases

**DOI:** 10.1101/2022.05.19.22275053

**Authors:** Lucy Findlater, Livia Pierotti, Charlie Turner, Adrian Wensley, Cong Chen, Shaun Seaman, Pantelis Samartsidis, Andre Charlett, Charlotte Anderson, Gareth Hughes, Matt Hickman, Obaghe Edeghere, Isabel Oliver

## Abstract

**Objective:** In September 2020, records of 15,861 SARS-CoV-2 cases failed to upload from the Second Generation Laboratory Surveillance System (SGSS) to the Contact Tracing Advisory Service (CTAS) tool, resulting in a delay in the contact tracing of these cases. This study used CTAS data to determine the impact of this delay on health outcomes: transmission events, hospitalisations, and mortality. Previously, a modelling study had suggested a substantial impact.

**Design:** Observational study

**Setting:** England.

**Population:** Individuals testing positive for SARS-CoV-2 and their reported contacts.

**Main outcome measures:** Secondary attack rates (SARs), hospitalisations, and deaths amongst primary and secondary contacts were calculated, compared to all other concurrent, unaffected cases. SGSS records affected by the event were matched to CTAS records and successive contacts and cases were identified.

**Results:** The initiation of contact tracing was delayed by 3 days on average in the primary cases in the delay group (6 days) compared to the control group (3 days). This was associated with lower completion of contact tracing of primary cases in the delay group: 80% (95%CI: 79-81%) in the delay group and 83% (95%CI: 83-84%) in the control group. There was some evidence to suggest an increase in transmission to non-household contacts amongst those affected by the delay. The SAR for non-household contacts was higher amongst secondary contacts in the delay group than the control group (delay group: 7.9%, 95%CI:6.4% to 9.2%; control group: 5.9%, 95%CI: 5.3% to 6.6%). There was no evidence of a difference between the delay and control groups in the odds of hospitalisation (crude odds ratio: 1.1 (95%CI: 0.9 to 1.2) or death (crude odds ratio: 0.7 (0.1 to 4.0)) amongst secondary contacts.

**Conclusions:** The delay in contact tracing had a limited impact on population health outcomes.

**Strengths and limitations of the study:**

- Shows empirical data on the health impact of an event leading to a delay in contact tracing so can test hypotheses generated by models of the potential impact of a delay in contact tracing
- Estimates the extent of further transmission and odds of increased mortality or hospitalisation in up to the third generation of cases affected by the event
- The event acts as a natural experiment to describe the possible impact of contact tracing, comparing a group affected by chance by delayed contact tracing to a control group who experienced no delay
- Contact tracing was not completed for all individuals, so the study might not capture all affected contacts or transmissions

## Introduction

As of April 2022, there have been over 21 million cases and 170,000 deaths from SARS-CoV-2 reported in the UK^1^. Contact tracing has been a central part of the public health response to SARS-CoV-2 and involves identifying contacts of people who have tested positive and advising them to self-isolate to reduce onward transmission of the virus^2^. On 28 May 2020, a national contact tracing system, NHS Test and Trace, was launched. All PCR positive cases in England were identified and and contacted by phone, digital tools, and through partnerships with local authorities^3 4^. Between 25 September and 2 October 2020, some data files containing test results from community testing sites failed to import from the laboratory surveillance system (Second Generation Surveillance System (SGSS)) to the contact tracing tool (Contact Tracing and Advisory Service (CTAS)). The files contained 15, 841 new positive cases^5^. At the time of the event, on the 28th September, there was a 7-day average of around 9500 reported cases per day in the UK with significant regional variation^1^.

The event did not affect the notification of test results to patients and did not affect the results of any specific testing sites, geographical areas, or groups of the population. However, the delay in the upload of case records to CTAS resulted in a delay in the contact tracing of affected cases, and therefore a potential delay in instructing their contacts to self-isolate, which could have resulted in further transmission of SARS-CoV-2, and therefore potentially more COVID-19 related hospitalisations and deaths. We expect that health outcomes of the initial group of cases (primary cases) whose records failed to upload to CTAS would not have been affected by the event, as they received their test results and advice to isolate in the usual timeframe, and transmission events from primary cases to their contacts (primary contacts who may become secondary cases) would have most likely occurred prior to the isolation of the case. However, these primary contacts may not have been aware that they were a close contact of a case or advised to isolate in a timely manner, which may have increased the risk of transmission to their contacts (secondary contacts who may become tertiary cases). The outcomes of secondary contacts are therefore the most likely to have been impacted by the delay in contact tracing. In addition, there could potentially be delays in secondary cases seeking healthcare support, increasing their risk of hospitalisation or excess mortality.

A published preliminary model suggested that this event may have been associated with an increase of up to a third additional infections and 30 - 40% additional deaths^6^.

In this study, we assess directly the health impact of the event, in terms of transmissions, hospitalisations, and deaths, using data on affected and concurrent cases and their contacts.

### Aims and objectives

The aims of this study are to:

- Describe the nature of any contact tracing delay experienced by cases affected by the event.
- Assess the extent of further transmission, by calculating attack rates from primary cases to their contacts and from secondary cases to their contacts.
- Assess the impact of the delay on health outcomes of primary and secondary contacts, in terms of hospitalisations and deaths.

## Methods

### Data sources

The list of SGSS records known to have been affected by the event was provided by the UK Health Security Agency (UKHSA). SGSS contains demographic and diagnostic information from laboratory test reports for patients who tested positive for SARS-CoV-2^7^. SGSS records were matched to COVID-19 testing and contact tracing records from the Contact Tracing Advisory Service (CTAS) database to validate the cases affected by the event. CTAS records represent SARS-CoV-2 case episodes, including information on the movements of cases in their infectious period, their contacts, and demographic and clinical characteristics (Supplementary Figure 1)^8^. Matching was conducted in repeated rounds based on combinations of the following identifiers: SGSS unique identifier; NHS number; forename; surname; date of birth; and postcode. This data set of CTAS records constituted the primary cases affected by the event, described henceforth as the ‘delay group’. A control group consisting of all primary cases from the same time window (30th September to 5th October 2020 inclusive) that were unaffected by the event was used as a comparison. The event affected exclusively cases arising from community testing sites and not hospital or other testing sites; therefore, primary cases in the control group were restricted to those arising from community testing^9^.

### Identifying cases and contacts

Contact records were linked to the case who reported them and assigned to the same group (delay or control group) as their associated case. Secondary cases were defined as (a) individuals reported as a contact by a primary case and (b) having a contact event with the primary case between 2 and 14 days inclusive prior to the onset of symptoms of the secondary case (or test date if no symptom onset available). If the secondary case was a household contact of the primary case, the date of interaction was taken to be the date of onset of symptoms (or test date if no symptom onset available) of the primary case. Secondary cases may be exposed to more than one primary case; therefore, only one transmission event was chosen to link a secondary case to the primary case who most likely infected them based on the following hierarchy: (1) a household transmission event was prioritised above all other types of contact event (due to the higher risk of transmission in this setting); (2) where multiple events of the same priority occurred, the most recent exposure was selected as the transmission event. Individuals who had taken multiple SARS-CoV-2 tests generated a CTAS record for each positive test result. These were identified by matching on name, DOB, NHS number, and contact details. For those cases, only the episode with the earliest test date was retained, as contact tracing should have been undertaken on receipt of the first positive result.

### Linkage to health outcomes

Contact data sets were linked to mortality data provided by UKHSA which describe death in people with a positive SARS-CoV-2 test. Matching was performed using NHS number. Two definitions of mortality consistent with SARS-CoV-2 surveillance were used: 1) death of a person with a laboratory-confirmed positive SARS-CoV-2 test and whose death was within 60 days of the first specimen date or more than 60 days after the first specimen date if COVID-19 is mentioned on the death certificate, and 2) death of a person with a laboratory-confirmed positive SARS-CoV-2 test and whose death was within (equal to or less than) 28 days of the first positive specimen date^10^. Contact datasets were also linked to the UKHSA hospital onset COVID-19 data set (extracted on 22 November 2021), which pulls from the identifiable daily feeds of data on hospitalisations: Secondary Uses Services (SUS+, a national data set describing patient hospitalisations); Emergency Care Data Set (ECDS, a national data set describing patient use of urgent and emergency care); and COVID-19 testing data. The data set provides one hospital spell per case if they had a hospital admission or discharge in the 14 day period before or after their first positive SARS-CoV-2 test, including people who were already in hospital at the time of test. Matching was performed using NHS number combined with date of birth, name, and sex.

### Descriptive analysis

Analysis was performed in R version 1.3.1056. Demographic information and contact tracing outcomes were described for the case and contact data sets (Supplementary Tables 2 and 3). Time taken for contact tracing was estimated using different dates in the contact tracing process: dates of test, laboratory report, SGSS record creation, CTAS record creation, and contact tracing completion (Supplementary Figure 2). Time taken to initiate the contact tracing process was defined as the days between the laboratory report date of the case and the date of upload of the case or contact record into CTAS. Time taken for the individual to complete contact tracing was defined as the days between the test date of the case and the date of contact tracing completion of the case or contact. Contact tracing completion was defined as the status of the individual’s CTAS record being set to ‘complete’ by a call handler or the individual submitting an online contact tracing form.

### Regression analysis

To understand the effect of the event on transmission, adjusted secondary attack rates (SARs) and differences in SARs amongst primary and secondary contacts were estimated via post-estimation from a logistic regression model. The predictor under investigation was whether the exposing case was in the delay or the control group. After review of crude SARs stratified by contact setting (household or non-household), effect modification of the predictor (delay or control group) by the contact setting (household or non-household) was allowed for in the model. The potential confounders included in the model were age group, sex, geographic region (one of nine UKHSA-defined geographical areas) of the contact, and whether the contact completed contact tracing.

To understand the effect of the event on health outcomes, odds ratios for experiencing mortality or hospitalisation amongst primary and secondary contacts were calculated using logistic regression with Firth penalisation, comparing individuals in the delay and control groups. When individuals had several contact episodes with multiple cases, the earliest episode per individual was retained. A very small number of mortality events was reported; therefore, unadjusted, crude odds ratios are presented to avoid overfitting the regression model, and the covariate balance between the delay and control groups suggests any major confounding is unlikely. As a sensitivity analysis, the health outcomes of the contacts in delay group were compared to a subgroup of contacts in the control group who were contact traced rapidly, defined as within three days of the date of test of the associated case.

### Patient and public involvement

Patients or the public were not involved in the design, or conduct, or reporting, or dissemination plans of this research.

## Results

### Sample characteristics

Overall, 15,861 SGSS records were identified as having been affected by the event and 15,467 (98%) were matched to a CTAS record (Supplementary Table 1). Following data cleaning, 15,285 (96%) primary cases affected by the delay remained eligible for inclusion in the study (Supplementary Figure 3). A control group of 43,742 concurrent primary cases was created, consisting of all CTAS records from the time period of 30^th^ September 2020 to 5^th^ October 2020 inclusive which were not affected by the event. Secondary and tertiary cases and primary and secondary contacts were identified in the CTAS database and demographic information is described in the supplementary material (Supplementary Tables 2 and 3).

### Nature of contact tracing delay

For primary cases in the delay group, it took on average two days longer for their records to be uploaded to CTAS, which initiated contact tracing, after the date the laboratory reported their positive test result (Table 1, Figure 1),. Primary cases also took on average three days longer to complete contact tracing in the delay group (6 days, interquartile range (IQR): 4-7) than the control group (3 days, (IQR: 2-5)) after their date of positive test. For primary contacts in the delay group, it took on average three days longer for their records to be uploaded to CTAS, relative to the laboratory report date of their associated case. It also took three days longer to complete contact tracing for primary contacts in the delay group (6 days (IQR: 5-8)) than the control group (3 days (IQR: 2-5)), relative to the test date of their associated case (Table 2, Figure 1). Once records had been uploaded onto CTAS, the primary cases and contacts in the delay group took the same median amount of time to complete contact tracing as the control group (1 day).

**Table 1:** Contact tracing outcomes for primary, secondary, and tertiary cases

|  | Delay group |  |  | Control group |  |  |
| --- | --- | --- | --- | --- | --- | --- |
| Cases | Primary | Secondary | Tertiary | Primary | Secondary | Tertiary |
| Number of observations | 15,285 | 2748 | 382 | 43,742 | 9575 | 1335 |
| Number of individuals | 15,285 (100.0%) | 2695 (98.1%) | 375 (98.2%) | 43,742 (100.0%) | 9382 (98.0%) | 1307 (97.9%) |
| Completed contact tracing (n (%; 95%CI)) |  |  |  |  |  |  |
| Yes | 12,221 (80.0%, 79.3%-81%) | 2137 (77.8%, 76.2%-79.3%) | 276 (72.3%, 67.5%-76.7%) | 36,503 (83.5%, 83.1%-83.8%) | 7291 (76.1%, 75.3%-77.0%) | 971 (72.7%, 70.3%-75.1%) |
| Difference in proportions who completed contact tracing (delay group minus control group) (95% CI) | -3.5% (-4.2% to -2.8%) | 1.6% (-0.2% to 3.4%) | -0.5% (-5.7% to 4.8%) | N/A | N/A | N/A |
| Median (and IQR) number of contacts reported per case |  |  |  |  |  |  |
| All | 3 (1-4) | 3 (1-4) | 3 (1-5) | 3 (1-4) | 3 (1-4) | 3 (1-4) |
| Household | 2 (1-3) | 2 (1-3) | 3 (1-4) | 2 (1-3) | 2 (1-4) | 2 (1-3) |
| Non-household | 0 (0-1) | 0 (0-0) | 0 (0-0) | 0 (0-1) | 0 (0-0) | 0 (0-0) |
| Mean number of contacts reported per case |  |  |  |  |  |  |
| All | 3.0 | 3.0 | 3.6 | 3.2 | 3.1 | 3.2 |
| Household | 2.2 | 2.4 | 2.6 | 2.3 | 2.5 | 2.4 |
| Non-household | 0.8 | 0.6 | 1.0 | 0.9 | 0.6 | 0.7 |
| Number of cases with ≥2 household contacts (n (%; 95%CI)) | 7361 (48.2%, 47.4%-49.0%) | 1348 (49.1%, 47.2%-50.9%) | 182 (47.6%, 42.5%-52.8%) | 22,244 (50.9%, 50.4%-51.3%) | 4717 (49.3%, 48.3%-50.3%) | 631 (47.3%, 44.6%-50.0%) |
| Median (and IQR) time taken for contact tracing, days between: |  |  |  |  |  |  |
| SGSS lab report date to CTAS upload date | 3 (2-4) | 1 (0-2) | 1 (0-1) | 1 (0-1) | 1 (0-1) | 1 (0-1) |
| Test date to CTAS completion date | 6 (4-7) | 4 (2-5) | 4 (2-5) | 3 (2-5) | 3 (2-5) | 3 (2-5) |
| Mean time taken for contact tracing, time between: |  |  |  |  |  |  |
| SGSS lab report date to CTAS upload date | 3.4 | 1.3 | 1.1 | 1.0 | 0.8 | 0.8 |
| Test date to CTAS completion date | 6.1 | 4.1 | 4.2 | 3.5 | 3.7 | 3.7 |

**Figure 1:**
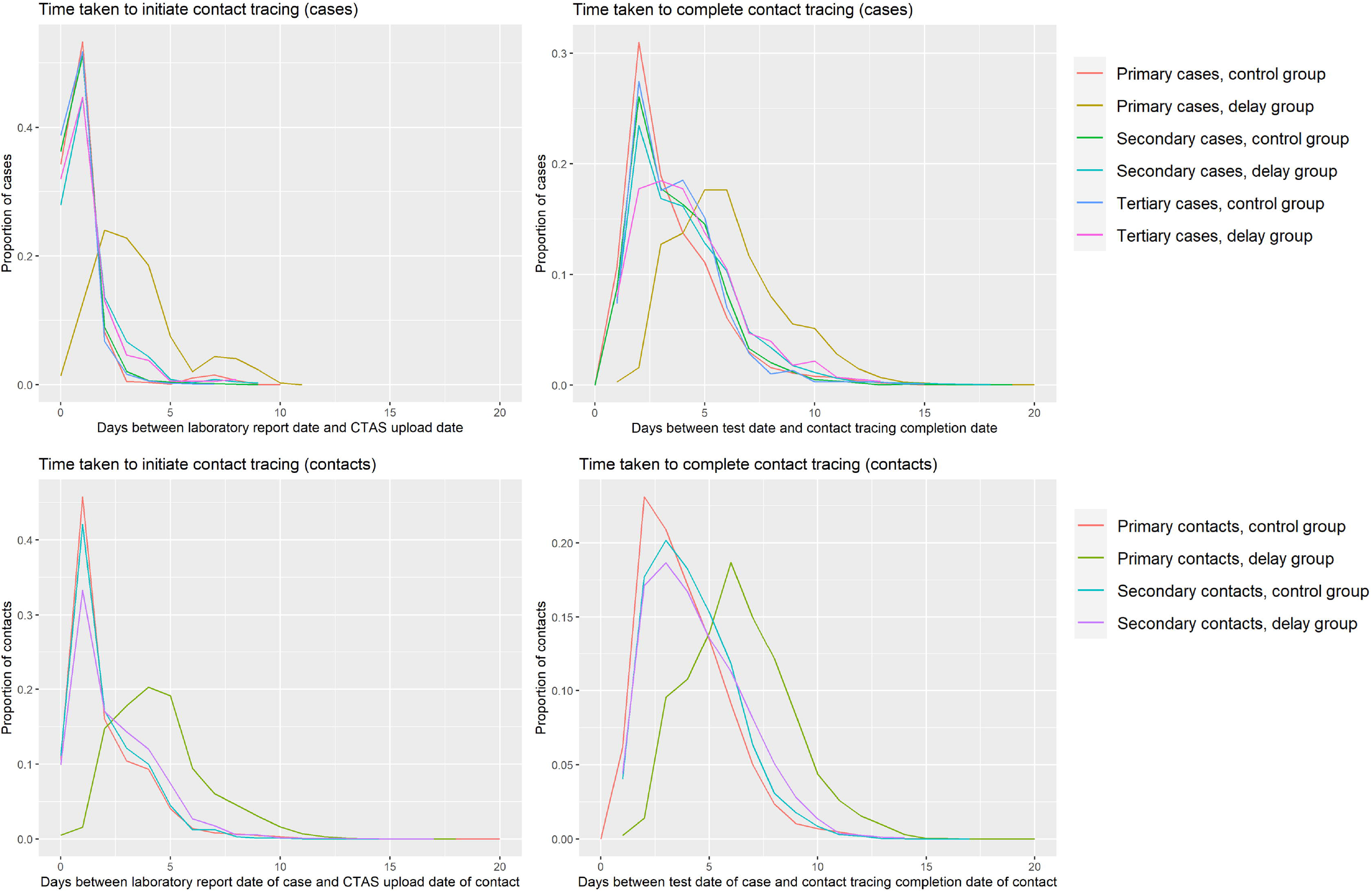
Graphs describing the time taken to initiate and complete contact tracing of cases and contacts in the delay and control groups

**Table 2:**
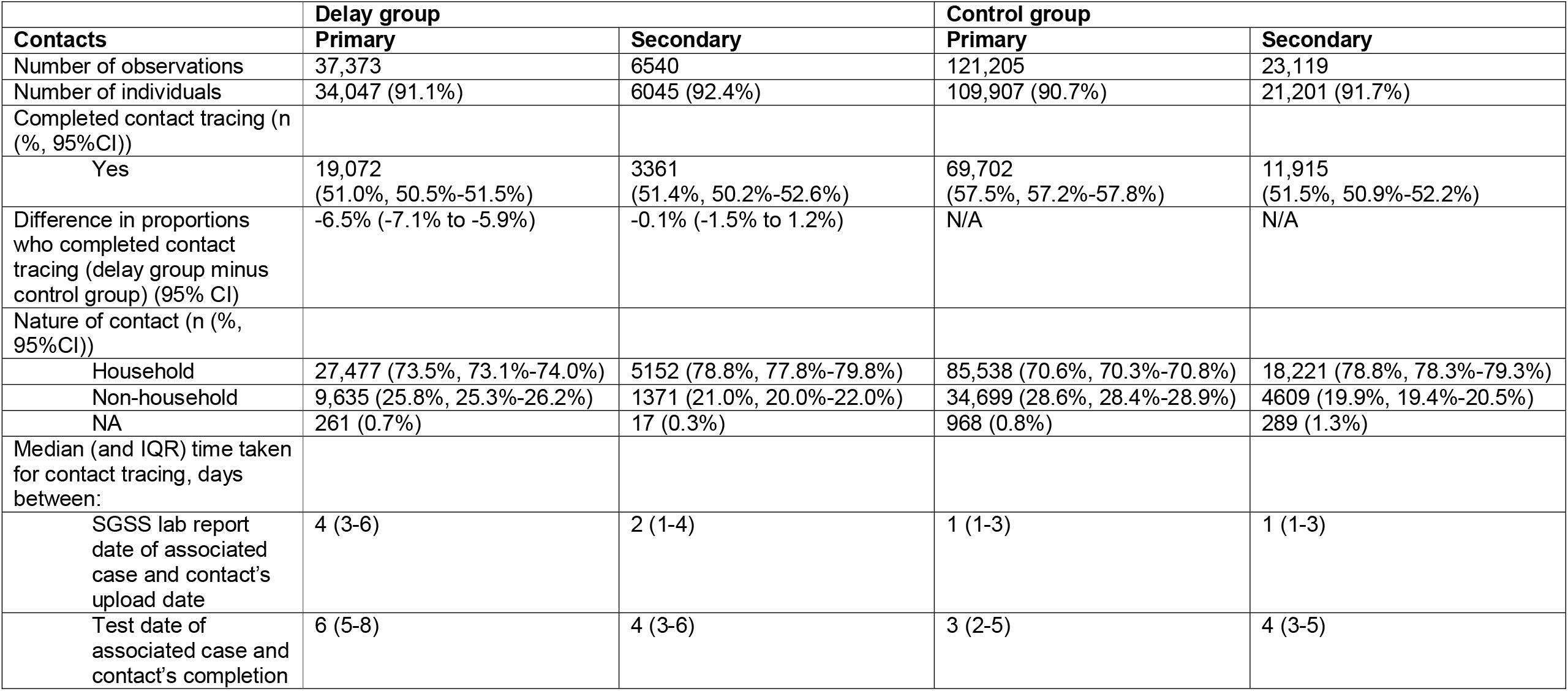
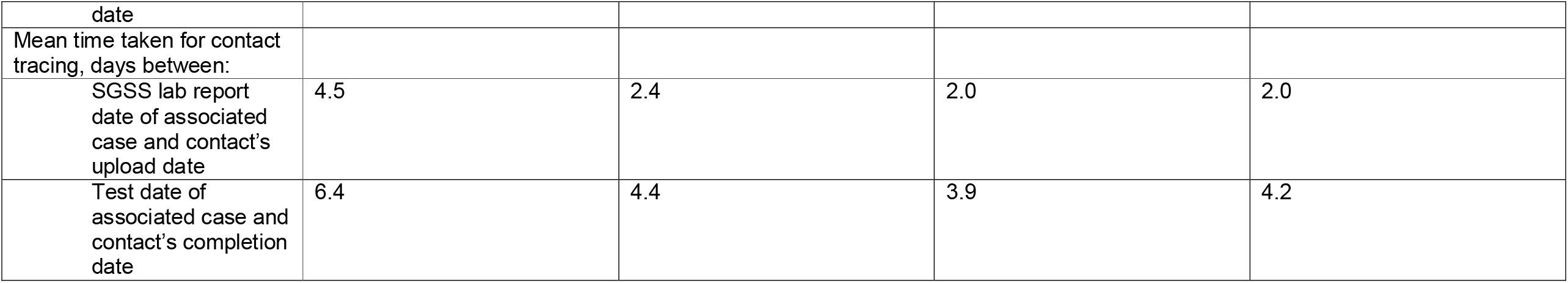
Contact tracing outcomes for primary and secondary contacts

|  | Delay group |  | Control group |  |
| --- | --- | --- | --- | --- |
| Contacts | Primary | Secondary | Primary | Secondary |
| Number of observations | 37,373 | 6540 | 121,205 | 23,119 |
| Number of individuals | 34,047 (91.1%) | 6045 (92.4%) | 109,907 (90.7%) | 21,201 (91.7%) |
| Completed contact tracing (n (%), 95%CI)) |  |  |  |  |
| Yes | 19,072 (51.0%, 50.5%-51.5%) | 3361 (51.4%, 50.2%-52.6%) | 69,702 (57.5%, 57.2%-57.8%) | 11,915 (51.5%, 50.9%-52.2%) |
| Difference in proportions who completed contact tracing (delay group minus control group) (95% CI) | -6.5% (-7.1% to -5.9%) | -0.1% (-1.5% to 1.2%) | N/A | N/A |
| Nature of contact (n (%), 95%CI)) |  |  |  |  |
| Household | 27,477 (73.5%, 73.1%-74.0%) | 5152 (78.8%, 77.8%-79.8%) | 85,538 (70.6%, 70.3%-70.8%) | 18,221 (78.8%, 78.3%-79.3%) |
| Non-household | 9,635 (25.8%, 25.3%-26.2%) | 1371 (21.0%, 20.0%-22.0%) | 34,699 (28.6%, 28.4%-28.9%) | 4609 (19.9%, 19.4%-20.5%) |
| NA | 261 (0.7%) | 17 (0.3%) | 968 (0.8%) | 289 (1.3%) |
| Median (and IQR) time taken for contact tracing, days between: |  |  |  |  |
| SGSS lab report date of associated case and contact's upload date | 4 (3-6) | 2 (1-4) | 1 (1-3) | 1 (1-3) |
| Test date of associated case and contact's completion | 6 (5-8) | 4 (3-6) | 3 (2-5) | 4 (3-5) |
| date |  |  |  |  |
| Mean time taken for contact tracing, days between: |  |  |  |  |
| SGSS lab report date of associated case and contact's upload date | 4.5 | 2.4 | 2.0 | 2.0 |
| Test date of associated case and contact's completion date | 6.4 | 4.4 | 3.9 | 4.2 |

The proportion of cases for whom contact tracing was completed was slightly lower for primary cases in the delay group (80%, 95%CI: 79-81%) than the control group (83%, 95%CI: 83-84%), a difference of -3.5% (95%CI: -4.2% to -2.8%) (Table 1). There was no difference in the proportion of secondary and tertiary cases who completed contact tracing. The proportion of primary contacts traced was lower by -6.5% (95%CI: -7.1% to -5.9%) in the delay group than the control group (delay group 51%, 95%CI: 51-52%; control group 58%, 95%CI: 57-58%), but there was no evidence of a difference for secondary contacts (delay group 51%, 95%CI: 50-53%; control group 52%, 95%CI: 51-52%) (Table 2). For primary and secondary cases, the median numbers of household and non-household contacts per case was the same in the delay and control groups; for tertiary cases, the median number was the same for non-household contacts but was greater by one person for household contacts in the delay compared to the control group (Table 1). For contacts, the nature of contact was reported as household contact for 71-79% of contacts in each group (Table 2).

### Effect on transmission

There was no evidence of a difference between the delay and control groups in the overall adjusted secondary attack rates (SARs) in primary or secondary contacts (Table 3). However, differences were observed in the SARs when stratified by household or non-household setting; SARs in non-household secondary contacts were higher for the delay group (7.9%, 95% CI 6.5% to 9.2%) than the control group (5.9%, 95% CI 5.3% to 6.6%), representing an increase of 1.9% (95% CI 0.4% to 3.4%). No difference was observed between the groups amongst household secondary contacts. Household SARs in primary contacts were lower for the delay group than for the control group. Conversely, for non-household contacts, the SARs in primary contacts were higher for the delay group (6.3%, 95% CI 5.8% to 6.8%) than for the control group (5.7%, 95% CI 5.5% to 5.9%), an increase of 0.6% (95% CI 0.1% to 1.1%).

**Table 3:** Adjusted secondary attack rates amongst contacts of primary and secondary cases in the delay group compared to the control group, by setting of contact

| Setting of contact event | Delay |  |  | Control |  |  | Difference in attack rates (95% CI)** |
| --- | --- | --- | --- | --- | --- | --- | --- |
|  | Number of contacts | Contacts becoming cases | Adjusted* secondary attack rate (SAR) (95% CI) | Number of contacts | Contacts becoming cases | Adjusted* secondary attack rate (SAR) (95% CI) |  |
| Primary contacts |  |  |  |  |  |  |  |
| All | 37,373 | 2,702 | 7.5% (7.3 to 7.8%) | 121,205 | 9,419 | 7.7% (7.5 to 7.8%) | -0.1% (-0.4 to 0.2%) |
| Household | 27,477 | 2,063 | 8.1% (7.8 to 8.4%) | 85,538 | 7,195 | 8.6% (8.4 to 8.7%) | -0.5% (-0.8 to -0.1%) |
| Non-household | 9,896 | 639 | 6.3% (5.8 to 6.8%) | 35,667 | 2,224 | 5.7% (5.5 to 5.9%) | 0.6% (0.1 to 1.1%) |
| Secondary contacts |  |  |  |  |  |  |  |
| All | 6,540 | 377 | 5.8% (5.2 to 6.3%) | 23,119 | 1,315 | 5.7% (5.4 to 6.0%) | 0.1% (-0.6 to 0.7%) |
| Household | 5,152 | 256 | 5.1% (4.5 to 5.7%) | 18,221 | 990 | 5.6% (5.3 to 6.0%) | -0.5% (-1.2 to 0.2%) |
| Non-household | 1,388 | 121 | 7.9% (6.5% to 9.2%) | 4,898 | 325 | 5.9% (5.3 to 6.6%) | 1.9% (0.4 to 3.4%) |

\*Adjusted SARs allowing for effect modification of the study type by household, and adjusted for the age group, sex and geographic region of the contact, and whether they completed contact tracing
\*\*Difference in attack rates calculated by subtracting control SAR from delay SAR

### Effect on health outcomes

COVID-19 death was a rare outcome in both groups with no evidence of a difference in 28 or 60 day mortality rates amongst all primary or secondary contacts in the delay group compared to the control group (Table 4). Using the 28 day definition of mortality, for primary contacts, there were 2.9 deaths (95%CI: 1.4 – 5.4) per 10,000 in the delay group compared to 3.7 deaths (95%CI: 2.7 – 5.1) per 10,000 in the control group. For secondary contacts, this was 0.0 (95%CI: 0.0 – 6.1) deaths per 10,000 in the delay group compared to 1.4 (95%CI: 0.3 – 4.1) deaths per 10,000 in the control group. There was no evidence for a difference observed in the odds of mortality for the delay group compared to the control group (Table 4, Figure 2).

**Figure 2:**
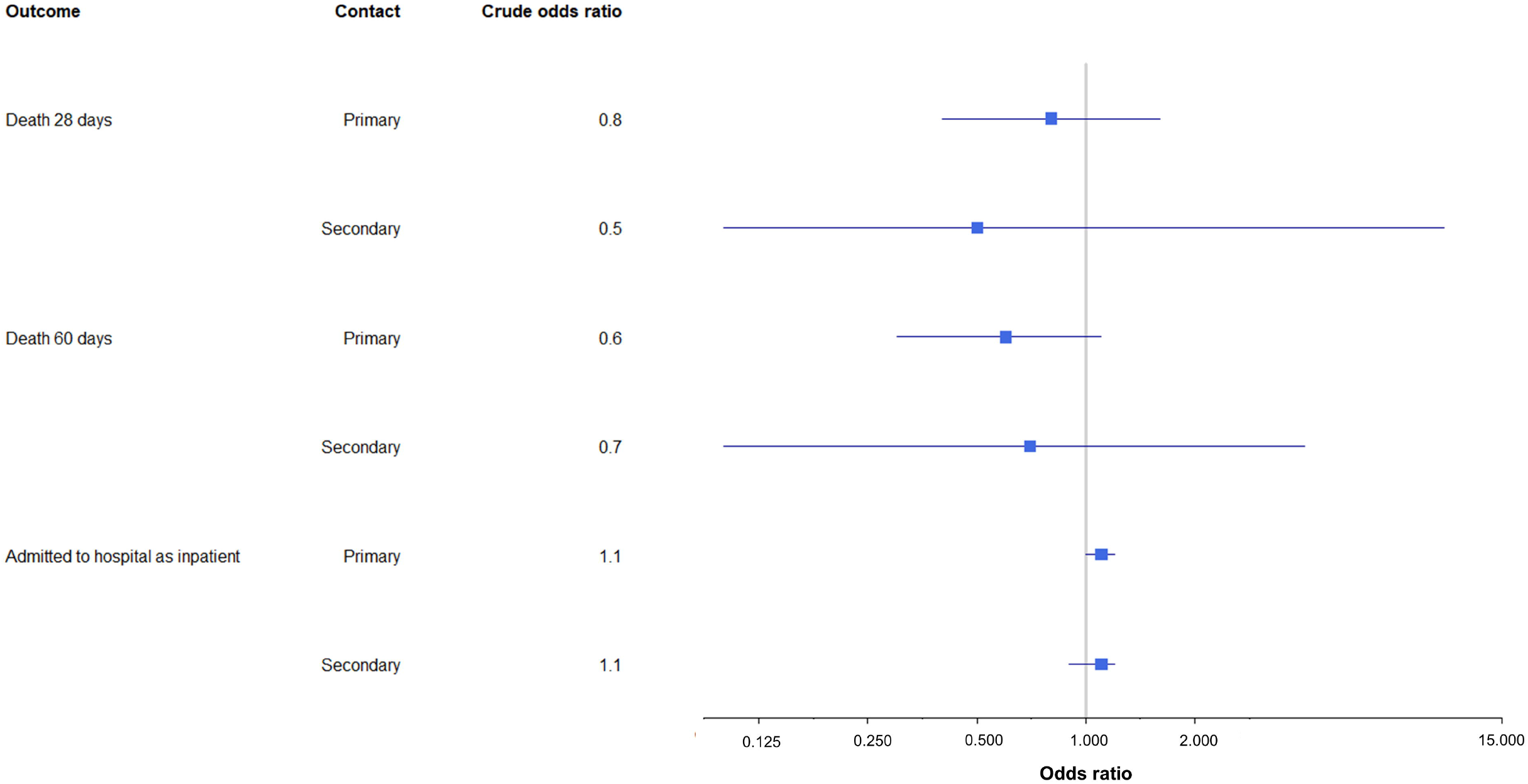
Graph describing odds ratios for mortality and hospitalisation, comparing the delay group to the control group (reference odds ratio = 1)

**Table 4:** Odds ratios for mortality and hospital outcomes in the delay group compared to the control group

|  | Delay group |  | Control group |  | Crude odds ratio* |
| --- | --- | --- | --- | --- | --- |
|  | Number (n = 34,047) | Proportion | Number (n = 109,907) | Proportion |  |
| **Primary contacts** |  |  |  |  |  |
| Deaths 28 day definition | 10 | 2.9 (1.4 – 5.4) per 10,000 population | 41 | 3.7 (2.7 – 5.1) per 10,000 population | 0.8 (0.4 to 1.6) |
| Deaths 60 day definition | 10 | 2.9 (1.4 – 5.4) per 10,000 population | 61 | 5.6 (4.2 – 7.1) per 10,000 population | 0.6 (0.3 to 1.1) |
| Admission to hospital as inpatient | 1121 | 3.3% (3.1% – 3.5%) | 3364 | 3.1% (3.0% - 3.2%) | 1.1 (1.0 to 1.2) |
| **Secondary contacts** | Number (n = 6045) | Proportion | Number (n = 21,201) | Proportion | Crude odds ratio* |
| Deaths 28 day | 0 | 0.0 (0.0 – 6.1) per 10,000 | 3 | 1.4 (0.3 – 4.1) per 10,000 | 0.5 (0.0 to 9.7) |
| definition |  | population |  | population |  |
| Deaths 60 day definition | 1 | 1.7 (0.0 – 9.2) per 10,000 population | 7 | 3.3 (1.3 – 6.8) per 10,000 population | 0.7 (0.1 to 4.0) |
| Admission to hospital as inpatient | 206 | 3.4% (3.0% – 3.9%) | 687 | 3.2% (3.0% – 3.5%) | 1.1 (0.9 to 1.2) |

\*The control group is the reference group (odds ratio = 1)

The proportion of contacts admitted to hospital as inpatients within 14 days of having their first COVID-19 test was almost identical in the delay and control groups, for both primary contacts (delay 3.3% (95%CI: 3.1-3.5%), control 3.1% (95%CI: 3.0-3.2%)) and secondary contacts (delay 3.4% (95%CI: 3.0-3.9%), control 3.2% (95%CI: 3.0-3.5%)) (Table 4), There was no significant difference in the crude or adjusted odds of hospital admission between the delay and control groups (Table 4, Figure 2). Additionally, there was no significant difference observed in the crude or adjusted odds of hospitalisation or death when the delay group was compared to a subset of the control group who were contact traced most rapidly (Supplementary Table 4).

## Discussion

### Statement of principal findings

The contact tracing of 15,861 SARS-CoV-2 cases affected by the event was delayed by an average of three days. There was evidence that the delay was associated with a slight decrease in the proportion of cases and contacts who completed contact tracing. Despite the delay, the mean and median number of overall contacts and non-household contacts made per case was similar in the delay group and the control group, for primary, secondary, and tertiary cases, and the median number of household contacts per case was near-identical.

As expected, we observed a higher SAR from secondary to tertiary cases, for non-household contact events in the delay group. There was no evidence of a difference in the SARs for secondary to tertiary cases overall or for household contact events. We found no evidence of a difference in hospitalisation or mortality among primary or secondary contacts in the delay or control group using the standard definitions for COVID-19-related hospitalisation and mortality used for surveillance purposes in the UK.

### Strengths and weaknesses

The event acted as a natural experiment whereby one group of individuals by chance experienced delayed contact tracing, enabling a comparison of their outcomes with concurrent, unaffected cases and contacts to assess the impact of contact tracing more generally. Having a a single national system collating all test results and a single national contact tracing system facilitated the study. Through record linkage we could identify successive generations of contacts and cases and describe key health outcomes associated with the delay in contact tracing; however, some of these outcomes were rare and it is possible that we did not have the power to detect a small difference in hospitalisation or mortality.

Secondary transmission could only be estimated using the contacts reported by cases who met the contact definition^11^. These would not include unknown contacts; however, the similarity in the number of reported contacts between the two groups suggests that unknown contacts are also likely to be similar. Contact tracing was not completed for a minority of cases in the delay and control groups, so there are likely to be further transmission events that are unknown and not described. People who do not engage in contact tracing differ from those who do in terms of ethnicity and socioeconomic status; however, this is unlikely to differ between the two groups^12 13 14^. Completion of contact tracing was slightly lower in the delay group. This could have potentially increased further the likelihood of worse outcomes in this group. It is important to note that there was an inherent lag in the initiation of contact tracing for all cases; therefore, the delay group was not compared to a perfect example of contact tracing, which may explain the lack of difference in observed health outcomes. To assess this, we conducted sensitivity analyses comparing the delay group to a subset of the control group who initiated contact tracing more quickly than the average for the control group, but we still did not detect a difference in health outcomes and the results were consistent with the main analysis. Some cases had multiple records, for example, if they had tested several times. For the analysis, we retained only the earliest CTAS episode. It is possible that some cases in the delay group reported subsequent positive tests soon after their first test which were not affected by the event (for example, after being admitted to hospital). Therefore, these people might still have been contact traced in a timely manner. Finally, primary cases were defined as being either affected by the event (delay group) or from the same time period (control group). Because the event occurred over a period of a small number of days, during which time transmission events occurred, primary cases generated some secondary and tertiary cases which were also classed as primary cases themselves and may have been directly affected by the event. Therefore, transmission chains may have been affected at different stages.

### Strengths and weaknesses in relation to other studies

A previous modelling exercise using aggregate published data projected substantial adverse health outcomes, describing a drastic rise in SARS-CoV-2 infections and deaths in areas most affected by the event and estimating over 125,000 additional infections and 1,500 additional COVID-19-related deaths^6^. Their rapid modelling approach described broader regional trends in infections and deaths using publicly available population surveillance data from the nearest calendar week to the event. This has the advantage of capturing all individuals possibly affected by the event, including any unknown or unreported contacts. However, it could lead to an overestimation of the health impact of the event. Our study described outcomes of the cases known to be affected by the event, which were a subset of the cases from 25 September – 2 October 2020, but the previous modelling study defined affected cases as all those with specimen dates from 20 – 27 September 2020; this might also lead to less accurate estimation of the health impact of the event. It is also important to note that the event occurred at a point in the pandemic when incidence of COVID-19 was increasing nationally and more rapidly in some regions^15 16^.

### Meaning of the study

Contacts of SARS-CoV-2 cases are at increased risk of infection, and this risk is higher among household contacts^17 18 19^. It is also known that significant numbers of infected people are asymptomatic and can transmit the virus^17 18 20^. Therefore, tracing all contacts of cases of COVID-19 to inform them of their exposure and give them advice on measures to reduce the risk of onwards transmission, has been a cornerstone of the pandemic response. This event offered an opportunity to assess the impact of contact tracing on health outcomes. Interestingly, few differences were observed between people impacted by the delay and other concurrent cases and contacts.

There was no difference in the number of contact events. This may reflect that people have generally maintained low levels of contacts throughout the pandemic^20^. We observed an increase in secondary transmission among non-household contacts but this was small. An increase in transmission in non-household contacts is to be expected if people are not aware of their exposure and therefore not isolating. We did not, however, observe any difference in SARs in household contacts, which may be explained by a more limited effect of self-isolation in preventing transmission within households. However, because many secondary cases were exposed to the virus in their household, their onward household attack rates could be lowered as their exposer would not become a case again within the time period studied.

Contact tracing is an effective tool to control transmission of infection^21 22 23^. Modelling studies have estimated the benefit from contact tracing and self-isolation to reduce SARS-CoV-2 transmission^24 25 26^. There is more limited evidence on the actual impact of the very large scale contact tracing undertaken during the pandemic. For contact tracing to be effective it needs to be timely and reach, as far as possible, all contacts^27 28 29^. The lack of a significant adverse health impact observed in our study could reflect the fact that there was an inherent delay in the tracing of COVID-19 cases. The lack of a difference may also reflect that, at the time, people generally were limiting their contact with others, with low numbers of contacts made by individuals in both groups^20^. Our study may also have lacked the power to detect a difference in the rare health outcome of mortality. Other factors that may have contributed include awareness of exposure status and need for self-isolation prior to official notification from the contact tracing system and a majority of contact events occurring in the household, where self-isolation is less likely to prevent transmission. Future research should evaluate the most effective approaches to conducting large scale contact tracing.

## Data Availability

The data that support this study were collected as part of a public health response and are considered sensitive and not made publicly available. Reasonable requests for access to anonymised data and data dictionary will be considered by the authors on request.

## Acknowledgements

We acknowledge the UK Health Security Agency (UKHSA) for permitting and facilitating access to data. We acknowledge Anne-Marie O’Connell, Richard Dunn, Tom Clare, Fernando Capelastegui, Russell Hope, Simon Collin, and Efejiro Ashano (UKHSA) and James Thomas (NHS Digital) for assisting with data acquisition and linkage. We acknowledge Daniela De Angelis (MRC Biostatistics Unit, UKHSA) for contributing to project development and feedback.

## Contributors

LF, LP, and IO contributed to conception and design of the work, analysis and interpretation of data, and drafted the manuscript. CT, AW, CC, CA, and GH contributed to design of the work, and acquisition, analysis, and interpretation of data. SS, PS, MH, and AC contributed to design of the work and interpretation of data. All authors revised the manuscript criticially and approved the final version of the manuscript.

## Disclaimer

The views expressed are those of the author and not necessarily those of the NIHR, the Department of Health and Social Care, or the UKHSA.

## Ethics approval

Approval for the study was granted from the Public Health England (PHE) (now UKHSA) Research Ethics and Governance Group (REGG). R&D reference: R&D 431.

## References

1 UK Government (2021). Coronavirus (COVID-19) in the UK. UK summary. Available from https://coronavirus.data.gov.uk/. Accessed on 25.04.22

2 Public Health England (2021) NHS Test and Trace Available from https://contact-tracing.phe.gov.uk/. Accessed on 01.12.21

3 Public Health England (2020) Letter from Richard Gleave, Deputy Chief Executive of Public Health England, describing the use of CTAS in the COVID-19 response. Available from https://assets.publishing.service.gov.uk/government/uploads/system/uploads/attachment_data/file/881231/LettertoDsPHoncontacttracing.pdf. Accessed on 01.12.21

4 Keeling et al. (2020) Efficacy of contact tracing for the containment of the 2019 novel coronavirus (COVID-19). J Epidemiol Community Health. 2020;74:861–866.

5 Public Health England (2020). PHE statement on delayed reporting of COVID-19 cases. Available from https://www.gov.uk/government/news/phe-statement-on-delayed-reporting-of-covid-19-cases. Accessed on 01.12.21

6 Fetzer and Graeber (2020). Does contact tracing work? Quasi-experimental evidence from an Excel error in England. CAGE working paper no. 521. Available from https://www.pnas.org/content/118/33/e2100814118. Accessed on 01.12.21

7 NHS Digital (2021). SGSS and CHESS data. Available from: https://digital.nhs.uk/about-nhs-digital/corporate-information-and-documents/directions-and-data-provision-notices/data-provision-notices-dpns/sgss-and-chess-data. Accessed on 01.12.21

8 UK Health Security Agency (2022). NHS Test and Trace in the workplace. Available from: https://www.gov.uk/guidance/nhs-test-and-trace-workplace-guidance. Accessed on 04.02.22

9 Department of Health & Social Care (2020). COVID-19 testing data: methodology note. Available from https://www.gov.uk/government/publications/coronavirus-covid-19-testing-data-methodology/covid-19-testing-data-methodology-note. Accessed on 01.12.21

10 Public Health England (2020). PHE data series on deaths in people with COVID-19: technical summary. Available from https://www.gov.uk/government/publications/phe-data-series-on-deaths-in-people-with-covid-19-technical-summary. Accessed on 01.12.21

11 UK Health Security Agency (2022). (Withdrawn) Guidance for contacts of people with confirmed coronavirus (COVID-19) infection who do not live with the person. Available from: https://www.gov.uk/government/publications/guidance-for-contacts-of-people-with-possible-or-confirmed-coronavirus-covid-19-infection-who-do-not-live-with-the-person/guidance-for-contacts-of-people-with-possible-or-confirmed-coronavirus-covid-19-infection-who-do-not-live-with-the-person. Accessed on 10.05.22

12 Shelby et al. (2021). Lessons learned from COVID-19 contact tracing during a public health emergency: a prospective implementation study. Frontiers in Public Health. 2021:9. doi: 10.3389/fpubh.2021.721952

13 Munzert S, Selb P, Gohdes A, et al. Tracking and promoting the usage of a COVID-19 contact tracing app. Nat Hum Behav 2021;5(2):247–55. doi: 10.1038/s41562-020-01044-x

14 Pegollo et al. (2021). Characteristics and determinants of population acceptance of COVID-19 digital contact tracing: a systematic review. Acta Biomedica. 2021;92(6):e2021444. doi: 10.23750/abm.v92iS6.12234

15 Office for National Statistics (2020). Coronavirus (COVID-19) Infection Survey pilot: England, Wales and Northern Ireland, 2 October 2020. Available from: https://www.ons.gov.uk/peoplepopulationandcommunity/healthandsocialcare/conditionsanddiseases/bulletins/coronaviruscovid19infectionsurveypilot/englandwalesandnorthernireland2october2020. Accessed on 11.05.21.

16 Office for National Statistics (2020). Coronavirus (COVID-19) Infection Survey pilot: England, Wales and Northern Ireland, 9 October 2020. Available from: https://www.ons.gov.uk/peoplepopulationandcommunity/healthandsocialcare/conditionsanddiseases/bulletins/coronaviruscovid19infectionsurveypilot/englandwalesandnorthernireland9october2020. Accessed on 11.05.21.

17 Marchant et al.(2021). Determining the acceptability of testing contacts of confirmed COVID-19 cases to improve secondary case ascertainment. J Public Health (Oxf). 2021;43(3):446–452. doi: 10.1093/pubmed/fdab079

18 Love et al. (2021). The acceptability of testing contacts of confirmed COVID-19 cases using serial, self-administered lateral flow devices as an alternative to self-isolation. medRxiv. doi: https://doi.org/10.1101/2021.03.23.21254168

19 UK Health Security Agency (2021). Insights on transmission of COVID-19 with a focus on the hospitality, retail and leisure sector. Available from: https://assets.publishing.service.gov.uk/government/uploads/system/uploads/attachment_data/file/982865/S1194_Transmission_in_hospitality_retail_leisure.pdf. Accessed on 01.12.21.

20 UK Health Security Agency (2021). Weekly statistics for NHS Test and Trace (England): 14 October to 20 October 2021. Available from: https://assets.publishing.service.gov.uk/government/uploads/system/uploads/attachment_data/file/1029536/test-and-trace-oct-14-to-20.pdf. Accessed on 01.12.21.

21 Hossain et al. (2022). Effectiveness of contact tracing in the control of infectious diseases: a systematic review. The Lancet Public Health. 2022;7(3):e259–e273

22 Bicher et al. (2021). Evaluation of Contact-Tracing Policies against the Spread of SARS-CoV-2 in Austria: An Agent-Based Simulation. Med Decis Making. 2021;41(8):1017–1032. doi: 10.1177/0272989X2110133062

23 Fields et al. (2021). Coronavirus Disease Contact Tracing Outcomes and Cost, Salt Lake County, Utah, USA, March–May 2020. Emerging Infectious Diseases. 27(12):2999–3008. doi: https://doi.org/10.3201/eid2712.210505

24 Kucharski et al (2020). Effectiveness of isolation, testing, contact tracing, and physical distancing on reducing transmission of SARS-CoV-2 in different settings: a mathematical modelling study. Lancet Infect Dis 2020;20(10):1151–1160. doi: 10.1016/S1473-3099(20)30457-6.

25 Chen and Huang (2022). Modelling the impacts of contact tracing on an epidemic with asymptomatic infection. Applied Mathematics and Computation. 2022;416. doi: https://doi.org/10.1016/j.amc.2021.126754

26 Amaku et al (2021). Modelling the impact of contact tracing of symptomatic individuals on the COVID-19 epidemic. Clinics (Sao Paulo). 2021;76:e2639. doi: 10.6061/clinics/2021/e2639. eCollection 2021nh

27 Ferretti et al. (2020). Quantifying SARS-CoV-2 transmission suggests epidemic control with digital contact tracing. Science. 2020;368(6491):eabb6936. doi: 10.1126/science.abb6936. Epub 2020 Mar 31.

28 Grantz et al. (2021). Maximizing and evaluating the impact of test-trace-isolate programs: A modelling study. PLOS Medicine. 2021;18(4): e1003585. doi: https://doi.org/10.1371/journal.pmed.1003585

29 Ashcroft P, Lehtinen S, Bonhoeffer S. Test-trace-isolate-quarantine (TTIQ) intervention strategies after symptomatic COVID-19 case identification. PloS one 2022;17(2):e0263597. doi: 10.1371/journal.pone.0263597

