## Supplementary Material for "Evaluating the impact on health outcomes of an event that resulted in a delay in contact tracing of COVID-19 cases"

**Supplementary Figure 1:** Definitions of SARS-CoV-2 cases and contacts

**Definitions of cases and contacts in the Contact Tracing Advisory Service (CTAS) database**

Individual records in the data represent case or contact episodes, where a case corresponds to a person with a positive test for SARS-CoV-2 and no case report in the 14 days prior. A contact is a person who was named by a case as being a contact between two days before the date of the case’s onset of symptoms (or test date, if not symptomatic) and the date of tracing. This includes being a member of or visitor to the case’s household, or close contact during an activity or in a work or education setting. An episode is an individual entry in NHS Test and Trace based on one of the above two definitions.

**Delay group**

**Primary case:** Confirmed case with laboratory evidence of infection as determined by detection of SARS-CoV-2 RNA by nucleic acid amplification, who had no case report in the 14 days prior to their test date, and who was known from SGSS records to have been impacted by the serious untoward incident (which affected some case records from 25 September to 2 October 2020).

**Primary contact**: Person who was identified as a contact of a primary case during their infectious period (between two days before the date of the case’s onset of symptoms or test date, if not symptomatic, and the date of tracing).

**Secondary case**: Person defined as a primary contact with evidence of infection as determined by detection of SARS-CoV-2 RNA by nucleic acid amplification and had a contact event with the primary case between two days and 14 days inclusive prior to the onset of symptoms (or test date if no symptom onset available) of the secondary case.

**Secondary contact**: Person who was identified as a contact of a secondary case during their infectious period (between two days before the date of the case’s onset of symptoms or test date, if not symptomatic, and the date of tracing).

**Tertiary case**: Person defined as a secondary contact with evidence of infection as determined by detection of SARS-CoV-2 RNA by nucleic acid amplification and had a contact event with the primary case between two days and 14 days inclusive prior to the onset of symptoms (or test date if no symptom onset available) of the tertiary case.

**Control group**

**Primary case:** Confirmed case with laboratory evidence of infection as determined by detection of SARS-CoV-2 RNA by nucleic acid amplification, who had no case report in the 14 days prior to their test date, and who was not on the list of case records affected by the incident, but had a CTAS upload date from within the time period of the incident (25 September to 2 October 2020 inclusive).

**Primary contact:** Person who was identified as a contact of a primary case during their infectious period (between two days before the date of the case’s onset of symptoms or test date, if not symptomatic, and the date of tracing).

**Secondary case:** Person defined as a primary contact with evidence of infection as determined by detection of SARS-CoV-2 RNA by nucleic acid amplification and had a contact event with the primary case between two days and 14 days inclusive prior to the onset of symptoms (or test date if no symptom onset available) of the secondary case.

**Secondary contact:** Person who was identified as a contact of a secondary case during their infectious period (between two days before the date of the case’s onset of symptoms or test date, if not symptomatic, and the date of tracing).

**Tertiary case:** Person defined as a secondary contact with evidence of infection as determined by detection of SARS-CoV-2 RNA by nucleic acid amplification and had a contact event with the primary case between two days and 14 days inclusive prior to the onset of symptoms (or test date if no symptom onset available) of the tertiary case.

**Supplementary Figure 2:** Diagram showing the different definitions for the time taken to initiate and complete contact tracing. The length of time taken for each stage in the diagram is the median time taken for that stage in the control group.

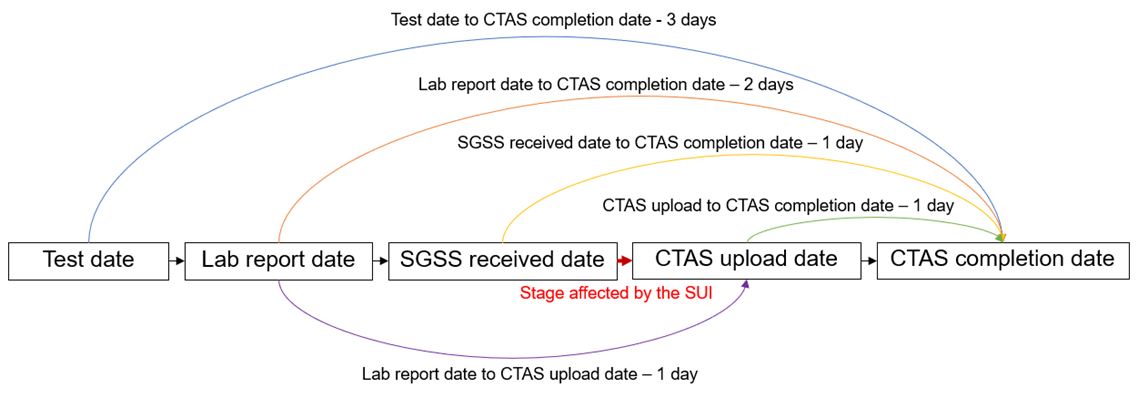

**Supplementary Table 1:** Description of each step of the process of matching SGSS records to CTAS records

| **Field(s) used for matching** | **Records matched (n = 15,861)** |
| --- | --- |
| 1. CDR Specimen Request SK number* | 14,655 |
| 2. NHS number, forename, surname, date of birth, postcode | 657 |
| 3. NHS number, forename, surname, date of birth | 64 |
| 4. NHS number, date of birth, postcode | 18 |
| 5. NHS number | 1 |
| 6. Forename, surname, date of birth, postcode | 72 |
| **Total matched** | **15,467 (98%)** |

* CDR Specimen Request SK number is the unique number identifier for each SGSS record

**Supplementary Figure 3**: Data flow chart describing exclusion criteria and de-duplication performed on the dataset of primary cases affected by the contact tracing delay caused by the incident. Only cases arising from community testing (pillar 2 testing) were affected; therefore, non-pillar 2 test episodes (including hospital testing) were excluded from the control group.

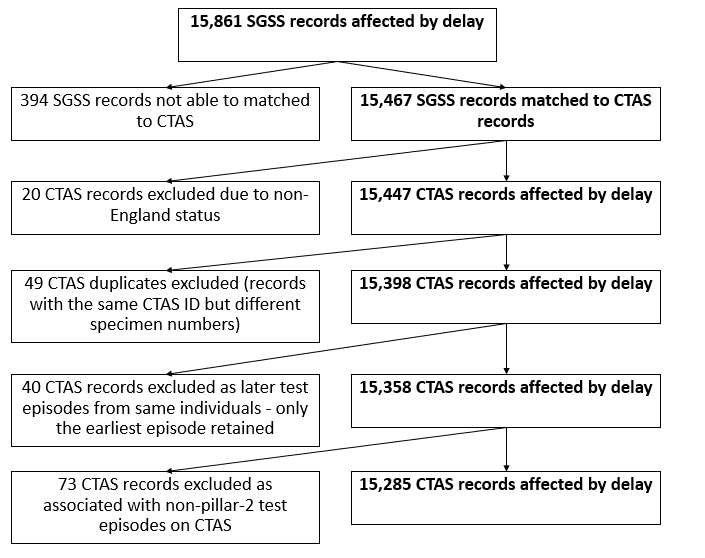

**Supplementary Table 2: Case demographics and contact tracing outcomes**

|  | **Delay group** | | | **Control group** | | |
| --- | --- | --- | --- | --- | --- | --- |
| **Cases** | **Primary** | **Secondary** | **Tertiary** | **Primary** | **Secondary** | **Tertiary** |
| Number of observations | 15,285 | 2748 | 382 | 43,742 | 9575 | 1335 |
| Number of individuals | 15,285 (100%) | 2695 (98%) | 375 (98%) | 43,742 (100%) | 9382 (98%) | 1307 (98%) |
| Sex |  |  |  |  |  |  |
| Male | 7300 (48%) | 1242 (45%) | 165 (43%) | 20,674 (47%) | 4186 (44%) | 606 (45%) |
| Female | 7985 (52%) | 1436 (52%) | 203 (53%) | 23,067 (53%) | 5105 (53%) | 692 (52%) |
| NA | 0 (0%) | 70 (3%) | 14 (4%) | 1 (0%) | 284 (3%) | 37 (3%) |
| Age (years) |  |  |  |  |  |  |
| < 18 | 1360 (9%) | 225 (8%) | 33 (9%) | 3509 (8%) | 881 (9%) | 158 (12%) |
| 18 – 29 | 6863 (45%) | 1303 (47%) | 198 (52%) | 19,806 (45%) | 4432 (46%) | 651 (49%) |
| 30 – 49 | 4030 (26%) | 591 (22%) | 75 (20%) | 10,951 (25%) | 2102 (22%) | 257 (19%) |
| 50 – 69 | 2585 (17%) | 556 (20%) | 70 (18%) | 7800 (18%) | 1852 (19%) | 234 (18%) |
| >= 70 | 447 (3%) | 68 (2%) | 6 (2%) | 1676 (4%) | 298 (3%) | 33 (3%) |
| NA | 0 (0%) | 5 (0%) | 0 (0%) | 0 (0%) | 10 (0%) | 2 (0%) |
| Ethnicity |  |  |  |  |  |  |
| Black/ African/ Caribbean/ Black British | 258 (2%) | 30 (1%) | 5 (1%) | 804 (2%) | 101 (1%) | 6 (0%) |
| Indian (Asian or Asian British) | 463 (3%) | 90 (3%) | 16 (4%) | 1420 (3%) | 296 (3%) | 38 (3%) |
| Mixed/ Multiple ethnic groups | 324 (2%) | 51 (2%) | 13 (3%) | 1071 (2%) | 181 (2%) | 22 (2%) |
| Other | 156 (1%) | 19 (1%) | 5 (1%) | 402 (1%) | 64 (1%) | 5 (0%) |
| Other Asian/ Asian British | 435 (3%) | 87 (3%) | 2 (1%) | 1179 (3%) | 192 (2%) | 24 (2%) |
| Pakistani (Asian or Asian British) | 702 (5%) | 91 (3%) | 13 (3%) | 1829 (4%) | 286 (3%) | 19 (1%) |
| White | 8525 (56%) | 1563 (57%) | 196 (51%) | 26,100 (60%) | 5466 (57%) | 748 (56%) |
| Not stated | 4422 (29%) | 817 (30%) | 132 (35%) | 10,937 (25%) | 2989 (31%) | 473 (35%) |
| Geographic region |  |  |  |  |  |  |
| East Midlands | 1042 (7%) | 202 (7%) | 40 (10%) | 3702 (8%) | 993 (10%) | 170 (13%) |
| East of England | 373 (2%) | 74 (3%) | 11 (3%) | 1234 (3%) | 290 (3%) | 55 (4%) |
| London | 1174 (8%) | 196 (7%) | 20 (5%) | 3966 (9%) | 755 (8%) | 111 (8%) |
| North East | 1974 (13%) | 338 (12%) | 50 (13%) | 4209 (10%) | 914 (10%) | 123 (9%) |
| North West | 5400 (35%) | 935 (34%) | 110 (29%) | 14,679 (34%) | 3125 (33%) | 372 (28%) |
| South East | 682 (4%) | 139 (5%) | 23 (6%) | 2385 (5%) | 489 (5%) | 91 (7%) |
| South West | 481 (3%) | 95 (3%) | 14 (4%) | 1718 (4%) | 336 (4%) | 55 (4%) |
| West Midlands | 1380 (9%) | 203 (7%) | 27 (7%) | 3707 (8%) | 791 (8%) | 115 (9%) |
| Yorkshire and the Humber | 2713 (18%) | 549 (20%) | 86 (23%) | 7876 (18%) | 1837 (19%) | 239 (18%) |
| NA | 66 (0%) | 17 (1%) | 1 (0%) | 266 (1%) | 45 (1%) | 4 (3%) |
| Completed contact tracing (n (%, 95%CI)) |  |  |  |  |  |  |
| Yes | 12221 (80%, 79-81%) | 2137 (78%, 76-79%) | 276 (72%, 67-77%) | 36,503 (83%, 83-84%) | 7291 (76%, 75-77%) | 971  (73%, 70-75%) |
| Difference in proportions who completed contact tracing (delay group minus control group) (95% CI) | -3.5% (-4.2% to -2.8%) | 1.6% (-0.2% to 3.4%) | -0.5% (-5.7% to 4.8%) | N/A | N/A | N/A |
| Median (and IQR) number of contacts reported per case |  |  |  |  |  |  |
| All | 3 (1-4) | 3 (1-4) | 3 (1-5) | 3 (1-4) | 3 (1-4) | 3 (1-4) |
| Household | 2 (1-3) | 2 (1-3) | 3 (1-4) | 2 (1-3) | 2 (1-4) | 2 (1-3) |
| Non-household | 0 (0-1) | 0 (0-0) | 0 (0-0) | 0 (0-1) | 0 (0-0) | 0 (0-0) |
| Mean number of contacts reported per case |  |  |  |  |  |  |
| All | 3.0 | 3.0 | 3.6 | 3.2 | 3.1 | 3.2 |
| Household | 2.2 | 2.4 | 2.6 | 2.3 | 2.5 | 2.4 |
| Non-household | 0.8 | 0.6 | 1.0 | 0.9 | 0.6 | 0.7 |
| Proportion of cases with ≥2 household contacts (n (%, 95%CI)) | 7361 (48%, 47-49%) | 1348 (49%, 47-51%) | 182 (48%, 43-53%) | 22,244 (51%, 50-51%) | 4717 (49%, 48-50%) | 631 (47%, 45-50%) |
| Median date of test | 28/09/2020 | 01/10/2020 | 03/10/2020 | 01/10/20 | 03/10/2020 | 05/10/2020 |
| Median (and IQR) time taken to initiate contact tracing, days between (median (IQR)): |  |  |  |  |  |  |
| Test date to CTAS completion date | 6 (4-7) | 4 (2-5) | 4 (2-5) | 3 (2-5) | 3 (2-5) | 3 (2-5) |
| SGSS lab report date to CTAS completion date | 4 (3-6) | 2 (1-4) | 2 (1-4) | 1 (1-3) | 2 (1-3) | 1 (1-3) |
| SGSS received date to CTAS completion date | 2 (1-3) | 1 (1-3) | 1 (1-3) | 1 (1-3) | 1 (1-3) | 1 (1-2) |
| CTAS upload date to CTAS completion date | 1 (1-2) | 1 (0-2) | 1 (0-2) | 1 (0-2) | 1 (0-2) | 1 (0-2) |
| SGSS lab report date to CTAS upload date | 3 (2-4) | 1 (0-2) | 1 (0-1) | 1 (0-1) | 1 (0-1) | 1 (0-1) |
| Mean time taken for contact tracing, time between: |  |  |  |  |  |  |
| SGSS lab report date to CTAS upload date | 3.4 | 1.3 | 1.1 | 1.0 | 0.8 | 0.8 |
| Test date to CTAS completion date | 6.1 | 4.1 | 4.2 | 3.5 | 3.7 | 3.7 |

**Supplementary Table 3: Contact demographics and contact tracing outcomes**

|  | **Delay group** | | **Control group** | |
| --- | --- | --- | --- | --- |
| **Contacts** | **Primary** | **Secondary** | **Primary** | **Secondary** |
| Number of observations | 37,373 | 6540 | 121,205 | 23,119 |
| Number of individuals | 34,047 (91%) | 6045 (92%) | 109,907 (91%) | 21,201 (92%) |
| Sex |  |  |  |  |
| Male | 8549 (23%) | 1486 (23%) | 31,318 (26%) | 5283 (23%) |
| Female | 10,002 (27%) | 1767 (27%) | 37,269 (31%) | 6260 (27%) |
| NA | 18,822 (50%) | 3287 (50%) | 52,618 (43%) | 11,576 (50%) |
| Age (years) |  |  |  |  |
| < 18 | 3000 (8%) | 394 (6%) | 9989 (8%) | 1653 (7%) |
| 18 – 29 | 7581 (20%) | 1548 (24%) | 30,151 (24%) | 5486 (24%) |
| 30 – 49 | 4319 (12%) | 695 (11%) | 15,050 (12%) | 2336 (10%) |
| 50 – 69 | 3480 (9%) | 599 (9%) | 12,464 (10%) | 2011 (9%) |
| >= 70 | 428 (1%) | 60 (1%) | 1875 (2%) | 246 (1%) |
| NA | 18,565 (50%) | 3244 (50%) | 51,706 (43%) | 11,387 (49%) |
| Ethnicity |  |  |  |  |
| Black/ African/ Caribbean/ Black British | 360 (1%) | 54 (1%) | 1352 (1%) | 178 (1%) |
| Indian (Asian or Asian British) | 722 (2%) | 147 (2%) | 2434 (2%) | 494 (2%) |
| Mixed/ Multiple ethnic groups | 558 (1%) | 90 (1%) | 2111 (2%) | 331 (1%) |
| Other | 190 (1%) | 30 (0%) | 636 (1%) | 78 (0%) |
| Other Asian/ Asian British | 641 (2%) | 124 (2%) | 2209 (2%) | 329 (1%) |
| Pakistani (Asian or Asian British) | 908 (2%) | 139 (2%) | 3107 (3%) | 428 (2%) |
| White | 14,236 (38%) | 2504 (38%) | 53,468 (44%) | 9122 (39%) |
| NA | 19,758 (53%) | 3452 (53%) | 55,888 (46%) | 12,159 (53%) |
| Geographic region |  |  |  |  |
| East Midlands | 2660 (7%) | 543 (8%) | 12,198 (10%) | 2836 (12%) |
| East of England | 932 (2%) | 163 (2%) | 3356 (3%) | 697 (3%) |
| London | 2469 (7%) | 436 (6%) | 9614 (8%) | 1742 (8%) |
| North East | 4949 (13%) | 828 (13%) | 11,818 (10%) | 2284 (10%) |
| North West | 12,753 (34%) | 2150 (33%) | 38,641 (32%) | 7064 (31%) |
| South East | 1599 (4%) | 324 (5%) | 6568 (5%) | 1087 (5%) |
| South West | 1194 (3%) | 229 (4%) | 5315 (4%) | 781 (3%) |
| West Midlands | 3405 (9%) | 486 (7%) | 10,175 (8%) | 2072 (9%) |
| Yorkshire and the Humber | 7293 (20%) | 1348 (21%) | 23,002 (19%) | 4444 (19%) |
| NA | 119 (0%) | 33 (1%) | 518 (0%) | 112 (0%) |
| Completed contact tracing (n (%, 95%CI)) |  |  |  |  |
| Yes | 19,072  (51%, 51-52%) | 3361 (51%, 50-53%) | 69,702  (58%, 57-58%) | 11,915 (52%, 51-52%) |
| Difference in proportions who completed contact tracing (delay group minus control group) (95% CI) | -6.5% (-7.1% to -5.9%) | -0.1% (-1.5% to 1.2%) | N/A | N/A |
| Nature of contact (n (%, 95%CI)) |  |  |  |  |
| Household | 27,477 (74%, 73-74%) | 5152 (79%, 78-80%) | 85,538 (71%, 70-71%) | 18,221 (79%, 78-79%) |
| Non-household | 9,635 (26%, 25-26%) | 1,371 (21%, 20-22%) | 34,699 (29%, 28-29%) | 4609 (20%, 19-20%) |
| NA | 261 (1%) | 17 (0%) | 968 (1%) | 289 (1%) |
| Median time taken to initiate contact tracing, days between (median (IQR)): |  |  |  |  |
| Test date of associated case and contact’s CTAS completion date | 6 (5-8) | 4 (3-6) | 3 (2-5) | 4 (3-5) |
| SGSS lab report date of associated case and contact’s completion date | 5 (4-7) | 3 (1-4) | 2 (1-4) | 2 (1-4) |
| SGSS received date of associated case and contact’s completion date | 3 (1-4) | 2 (1-4) | 2 (1-3) | 2 (1-3) |
| CTAS upload date of case and contact’s completion date | 2 (1-3) | 2 (1-3) | 1 (0-3) | 2 (1-3) |
| CTAS upload date of contact and contact’s completion date | 1 (0-2) | 1 (0-2) | 1 (0-1) | 1 (0-1) |
| SGSS lab report date of associated case and contact’s CTAS upload date | 4 (3-6) | 2 (1-4) | 1 (1-3) | 1 (1-3) |
| Mean time taken for contact tracing, days between |  |  |  |  |
| SGSS lab report date of associated case and contact’s upload date | 4.5 | 2.4 | 2.0 | 2.0 |
| Test date of associated case and contact’s completion date | 6.4 | 4.4 | 3.9 | 4.2 |
| Estimated compliance of cases with self-isolation, for non-household contact events only* |  |  |  |  |
| Contact event date was before test date of associated case | 9276 (94%) (n = 9820) | 1289 (93%) (n = 1383) | 33,284 (94%) (n = 35,368) | 4689 (97%) (n = 4852) |
| Contact event date was before laboratory report date of associated case | 9553 (97%) (n = 9816) | 1335 (97%) (n = 1377) | 34,867 (99%) (n = 35,277) | 4744 (98%) (n = 4822) |
| Contact event date was before symptom onset date of associated case | 7245 (78%) (n = 9267) | 1069 (83%) (n = 1288) | 25,167 (76%) (n = 33,136) | 3843 (87%) (n = 4437) |

*The compliance of cases with self-isolation was explored by describing whether the date of test, laboratory report, and symptom onset of the case occurred before or after the date of contact with their associated contacts, for non-household contacts only. If the case made non-household contacts after taking a test, developing symptoms, or being informed they had tested positive (using laboratory report date as a proxy), this suggests a lack of compliance with self-isolation.

**Supplementary Table 4:** Description of the odds ratio for primary and secondary contacts of experiencing health outcomes (mortality or hospitalisation) comparing the delay group to (1) all controls in the control group and (2) to rapidly traced controls in the control group*

|  | **Comparison of odds in each group*** |
| --- | --- |
| **Primary contacts** | **Crude odds ratio** |
| Deaths 28 day definition |  |
| All controls | 0.9 (0.4 to 1.6) |
| Rapidly traced controls | 0.7 (0.3 to 1.3) |
| Deaths 60 day definition |  |
| All controls | 0.6 (0.3 to 1.1) |
| Rapidly traced controls | 0.5 (0.2 to 1.0) |
| Admission to hospital as inpatient |  |
| All controls | 1.1 (1.0 to 1.2) |
| Rapidly traced controls | 1.1 (1.0 to 1.1) |
| **Secondary contacts** |  |
| Deaths 28 day definition |  |
| All controls | 0 (NA to Inf) |
| Rapidly traced controls | 0 (NA to Inf) |
| Deaths 60 day definition |  |
| All controls | 0.5 (0.0 to 2.8) |
| Rapidly traced controls | 0.5 (0.0 to 3.4) |
| Admission to hospital as inpatient |  |
| All controls | 1.1 (0.9 to 1.3) |
| Rapidly traced controls | 0.9 (0.8 to 1.1) |

*The control group is the reference group (odds ratio = 1), compared to the delay group. The control group consisted of either all controls, or rapidly traced controls, defined as contacts with a date of contact tracing completion within 3 days of the date of test of the associated case.
